# A metabolome-wide Mendelian randomization study prioritizes causal circulating metabolites for multiple sclerosis

**DOI:** 10.1101/2022.11.11.22282226

**Authors:** Angela Ge, Yitang Sun, Thaddaeus Kiker, Yanjiao Zhou, Kaixiong Ye

## Abstract

**Objective:** To prioritize circulating metabolites that likely play causal roles in the development of multiple sclerosis (MS).

**Methods:** Two-sample Mendelian randomization analysis was performed to estimate the causal effects of 571 circulating metabolites on the risk of MS. Genetic instruments for circulating metabolites were obtained from three previous genome-wide association studies (GWAS) of the blood metabolome, while genetic associations with MS were from a large GWAS by the International Multiple Sclerosis Genetics Consortium. The primary analysis was performed with the multiplicative random-effect inverse variance-weighted method, while multiple sensitivity analyses were conducted with the weighted median, weighted mode, MR-Egger, and MR-PRESSO.

**Results:** A total of 29 metabolites had suggestive evidence of causal associations with MS. Genetically instrumented levels of serine (OR = 1.56, 95% CI = 1.25 – 1.95), lysine (OR = 1.18, 95% CI = 1.01 – 1.38), acetone (OR = 2.45, 95% CI = 1.02 – 5.90), and acetoacetate (OR = 2.47, 95% CI = 1.14 – 5.34) were associated with a higher MS risk. Total cholesterol and phospholipids in large very-low-density lipoprotein were associated with a lower MS risk (OR = 0.83, 95% CI = 0.69 – 1.00; OR = 0.80, 95% CI = 0.68 – 0.95), but risk-increasing associations (OR = 1.20, 95% CI = 1.04 – 1.40; OR = 1.13, 95% CI = 1.00 – 1.28) were observed for the same two lipids in very large high-density lipoprotein.

**Conclusions:** Our metabolome-wide Mendelian randomization study prioritized a short list of circulating metabolites, such as serine, lysine, acetone, acetoacetate, and lipids, that likely have causal associations with MS.

## Introduction

Multiple sclerosis (MS) is an autoimmune disease in the central nervous system, characterized by demyelination and neurodegeneration. While the exact causes of MS are still unknown, some lifestyle and environmental risk factors have been relatively well-established, such as female sex, smoking, Epstein–Barr virus (EBV) infection, low vitamin levels, and obesity^1^. Metabolomics is a powerful approach to identifying metabolites that differentiate MS patients from healthy controls, revealing diagnostic or prognostic biomarkers, potential therapeutic targets, and insights into the pathogenesis^2, 3^. Metabolites in various metabolic pathways have been implicated in MS, such as higher plasma levels of acetoacetate, acetone, and 3-hydroxybutyrate in energy metabolism^4^, higher circulating levels of gamma-glutamyl amino acids and lysine in amino acid metabolism^5, 6^, elevated serum levels of uridine in nucleotide metabolism^7^, and altered circulating profiles of lipids and lipoproteins in lipid metabolism^8^. However, the causality of these lifestyle, environmental or metabolomic risk factors is hard to establish due to the inherent limitations of observational associations, especially in case-control studies, such as reverse causation and residual confounding^1^.

Mendelian randomization (MR) is a genetic epidemiology method that leverages genetic effects to enable the inference of causality between an exposure and an outcome. It selects genetic variants with known effects on the exposure of interest. The random allocation of the two alleles at a genetic variant across generations mimics the random assignment of placebo or treatment to participants in a randomized controlled trial^9^. MR has been applied to MS, providing evidence for causal roles of high BMI^10-13^, increased interleukin-6 (IL-6) signaling^11^, and low vitamin D^10, 13^. On the other hand, MR did not support the causal roles of uric acid^14^, leptin^13^, adiponectin^13^, and depression^15, 16^. A systemic MR study of 65 possible risk factors for MS revealed robust evidence of causality for four of them, high childhood and adult BMI, low vitamin D, and low physical activity. It also found suggestive evidence for type 2 diabetes, waist circumference, body fat percentage, age of puberty, and high-density lipoprotein cholesterol (HDL-C)^17^. Although large-scale MR analysis is a powerful approach to prioritizing causal risk factors for MS, no such study has been applied to all metabolites in a metabolome. Taking advantage of the recent large genome-wide association studies (GWAS) of human blood metabolites measured by metabolomics platforms^18-20^, we performed a metabolome-wide MR study to prioritize causal circulating metabolites for MS.

## Methods

### Data Sources

Genetic associations with circulating metabolites were obtained from three GWAS of human blood metabolome. Their summary statistics were compiled and made available through the MRC IEU OpenGWAS project^21, 22^, and the three GWAS were labeled as met-a^18^, met-c^19^, and met-d^20^. All three GWAS were performed in participants of European ancestry. The met-a study covers 452 metabolites and 7,824 participants, the met-c study 123 metabolites and up to 24,925 participants, and the met-d study 249 metabolites and 115,078 individuals. For met-a, which performed GWAS on raw phenotypes, we rescaled SNP effect sizes to one standard deviation (SD) of the circulating metabolite level^18^. The effect sizes in met-c and met-d were already standardized to SD because of the inverse rank-based normal transformation of phenotypic values before GWAS^19, 20^. Genetic associations with MS in Europeans were obtained from the discovery GWAS by the International Multiple Sclerosis Genetics Consortium (14,802 cases and 26,703 control). We obtained access to the summary statistics on the designated website (https://nettskjema.no/a/imsgc-data-access#/). We further confirmed that the same summary statistics were available on the OpenGWAS project with a dataset ID of ieu-b-18.

### Selection of Instrumental Variables

Two significance thresholds (*P* < 5 × 10^−8^ and *P* < 1 × 10^−6^) were used to select single nucleotide polymorphisms (SNPs) as instrumental variables (IV). This enables the inclusion of more metabolites in our MR analysis and the evaluation of the robustness of the effect estimates. We used linkage disequilibrium (LD) clumping (*r*^2^ < 0.001 within a 10 Mb window) to identify independent SNPs. For exposure-associated SNPs not present in the MS GWAS dataset, we searched for proxy SNPs in high LD (*r*^2^ ≥ 0.8). A threshold of F-statistics greater than 10 indicates strong instruments, avoiding weak instrument bias^23^. The effects of IVs on exposure and outcome were harmonized to rule out strand mismatches and ensure alignment of effect sizes. All IV selection, clumping, and harmonization were implemented in R v4.2.1 using the TwoSampleMR package (v0.5.6)^21^.

### Statistical Analyses

We performed two-sample MR analysis only for metabolites that have at least three independent genetic instruments in order to apply statistical testing of and correction for potential pleiotropy. The multiplicative random-effect inverse variance-weighted (IVW) method was our primary method^24^. The Cochran’s Q test was used to determine the homogeneity within the causal estimates of different SNPs^25^. Sensitivity analyses were performed with MR-Egger^26^, weighted median (WME)^27^, and weighted mode (WMO) methods^28^. The MR-Egger intercept test was applied to evaluate the presence of unbalanced horizontal pleiotropy^26, 29^. Moreover, we applied the MR-PRESSO method for detecting overall horizontal pleiotropy (i.e., the global test), identifying specific outliers (i.e., the outlier test), and re-calculating effect estimates after outlier removal^30^. In addition, we applied the MR Steiger method to infer the direction of causality^31^. All analyses were conducted in R v4.2.1 using MendelianRandomization (v0.6.0)^32^, TwoSampleMR (v0.5.6)^21^, and MRPRESSO (v1.0)^30^.

### Standard protocol approvals, registrations, and patient consent

This study was conducted with previously published summary-level data. No individual-level data were used.

### Data availability

All GWAS summary statistics could be accessed through the OpenGWAS project^21, 22^. The MR analysis scripts can be found at https://github.com/yitangsun/metabolome-wide-MR-for-MS.

## Results

Our workflow is summarized in Figure 1. Three GWAS of human blood metabolome were included in our analysis, each with 452, 123, and 249 metabolites, respectively. Metabolites with less than three genetic instruments were excluded from our analysis. With a significance cutoff of *P* < 5 × 10^−8^ for the selection of genetic instruments, we obtained MR results for a total of 404 metabolites (Supplementary Table 1). When we relaxed the significance cutoff to *P* < 1 × 10^−6^, an additional 167 metabolites were included for MR analysis, reaching a total of 571 metabolites (Supplementary Table 2). For all the 404 metabolites included in analyses with both significance cutoffs, the effect estimates are highly concordant (Supplementary Figure 1). A total of 29 metabolites have supporting evidence for a causal role in MS. Six metabolites have nominally significant and directionally consistent effect estimates with both significance cutoffs. Another 23 metabolites have nominally significant signals with one cutoff, and the effect estimates are close to those based on the other cutoff, when available (Figure 2). Sensitivity analyses with MR-Egger, WME, WMO, and MR-PRESSO revealed directionally consistent effect estimates, although not always reaching nominal significance. The MR Steiger test further supports the causal direction from the metabolite to MS, instead of the reverse (Supplementary Tables 1 and 2).

**Figure 1.**
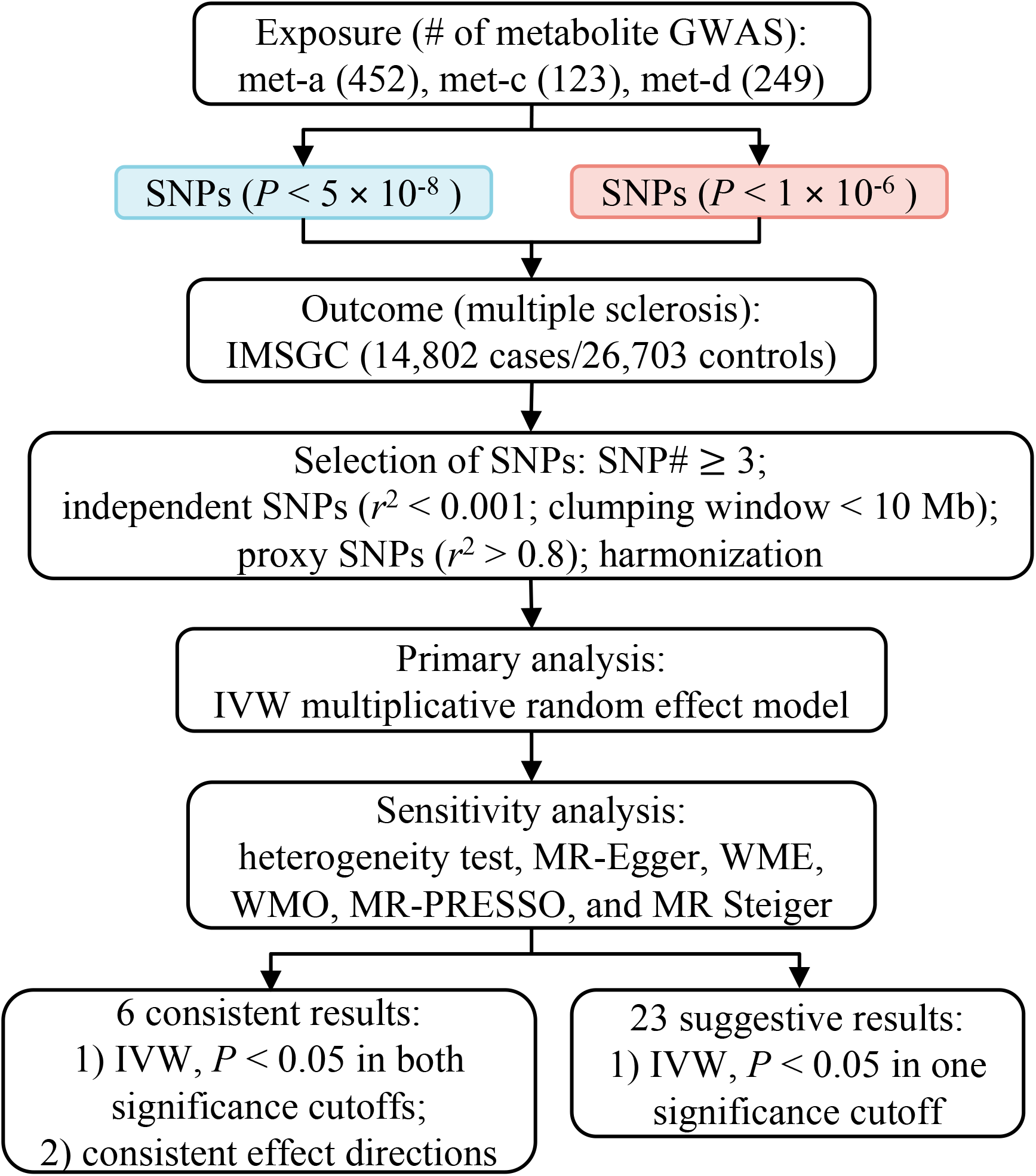
Flowchart of the MR study. MR: Mendelian randomization; GWAS: genome-wide association studies; SNPs: single nucleotide polymorphisms; IMSGC: International Multiple Sclerosis Genetics Consortium; IVW: inverse variance-weighted; WME: weighted median; WMO: weighted mode.

**Figure 2.**
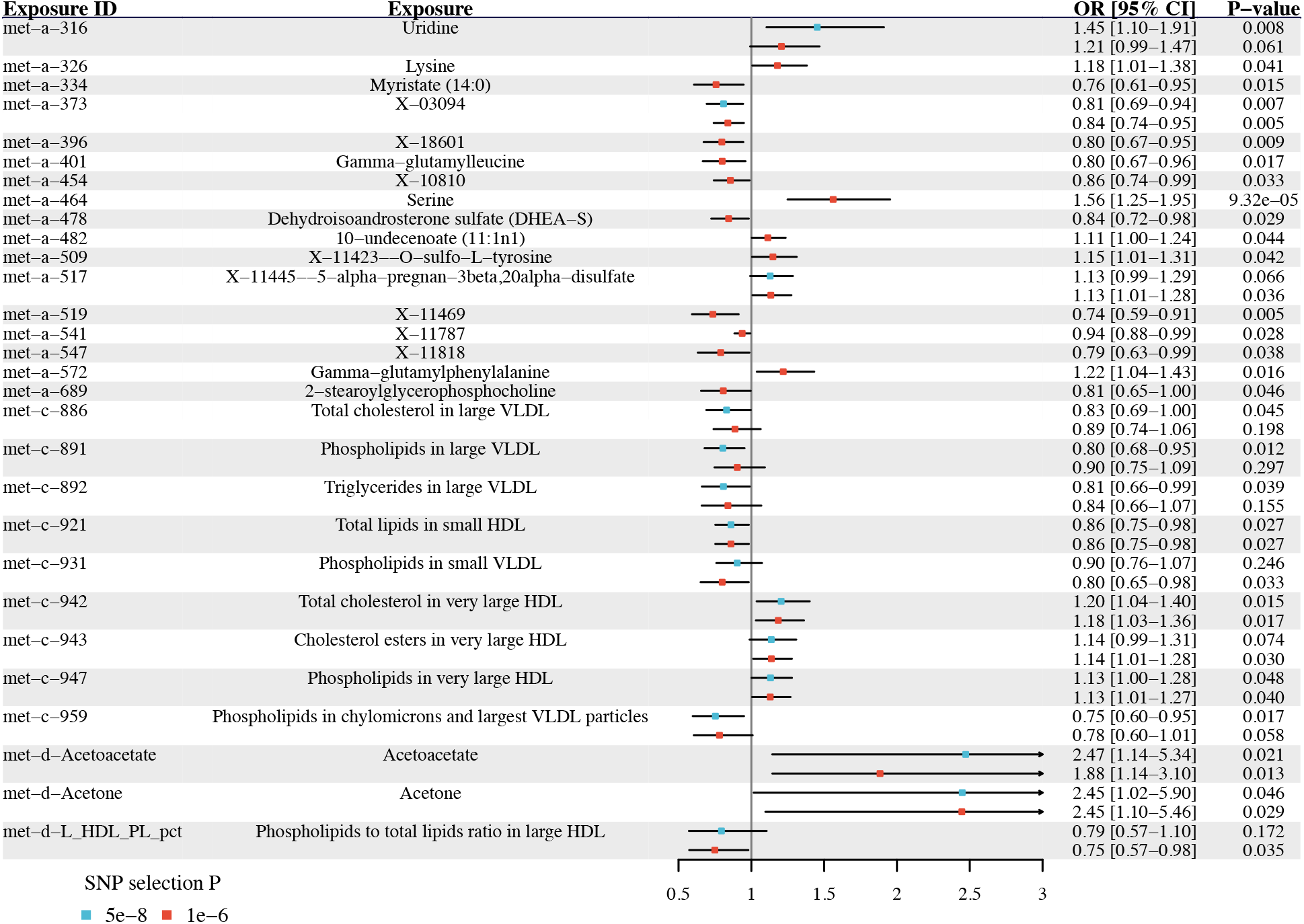
Metabolites with significant MR estimated effects on the risk of MS. Odds ratios and 95% confidence intervals are scaled to per genetically predicted 1 SD increase in circulating metabolite levels. MR: Mendelian randomization; MS: multiple sclerosis; OR: odds ratios; 95% CI: 95% confidence intervals; SD: standard deviation; SNP: single nucleotide polymorphism.

Among the 29 metabolites with supporting evidence, ten are lipids in specific lipoprotein subclasses. The genetically predicted circulating levels of total cholesterol (OR = 0.83, 95% CI = 0.69 – 1.00, *P* = 0.045), phospholipids (OR = 0.80, 95% CI = 0.68 – 0.95, *P* = 0.012), and triglycerides (OR = 0.81, 95% CI = 0.66 – 0.99, *P* = 0.039) in large very-low-density lipoprotein (VLDL) are associated with a lower risk of MS. Phospholipids in small VLDL (OR = 0.80, 95% CI = 0.60 – 0.95, *P* = 0.017) and in chylomicrons and the largest VLDL particles (OR = 0.75, 95% CI = 0.65 – 0.98, *P* = 0.033) are both negatively associated with the MS risk. In contrast, the genetically predicted circulating levels of total cholesterol (OR = 1.20, 95% CI = 1.04 – 1.40, *P* = 0.015), phospholipids (OR = 1.13, 95% CI = 1.00 – 1.28, *P* = 0.048), and cholesterol esters (OR = 1.14, 95% CI = 1.01 – 1.28, *P* = 0.030) in very large HDL are positively associated with the risk of MS.

Five of the 29 metabolites are amino acids. The genetically predicted circulating levels of serine (Figure 3A and 3D, OR = 1.56, 95% CI = 1.25 – 1.95, *P* = 9.32 × 10^−5^), lysine (OR = 1.18, 95% CI = 1.01 – 1.38, *P* = 0.041) and O-sulfo-L-tyrosine (OR = 1.15, 95% CI = 1.01 – 1.31, *P* = 0.042) are all associated with a higher risk of MS. The other two are gamma-glutamyl amino acids, and they have opposite associations. Gamma-glutamyl leucine is negatively (OR = 0.80, 95% CI = 0.67 – 0.96, *P* = 0.017), while gamma-glutamylphenylalanine is positively (OR = 1.22, 95% CI = 1.04 – 1.43, *P* = 0.016) associated with the MS risk.

**Figure 3.**
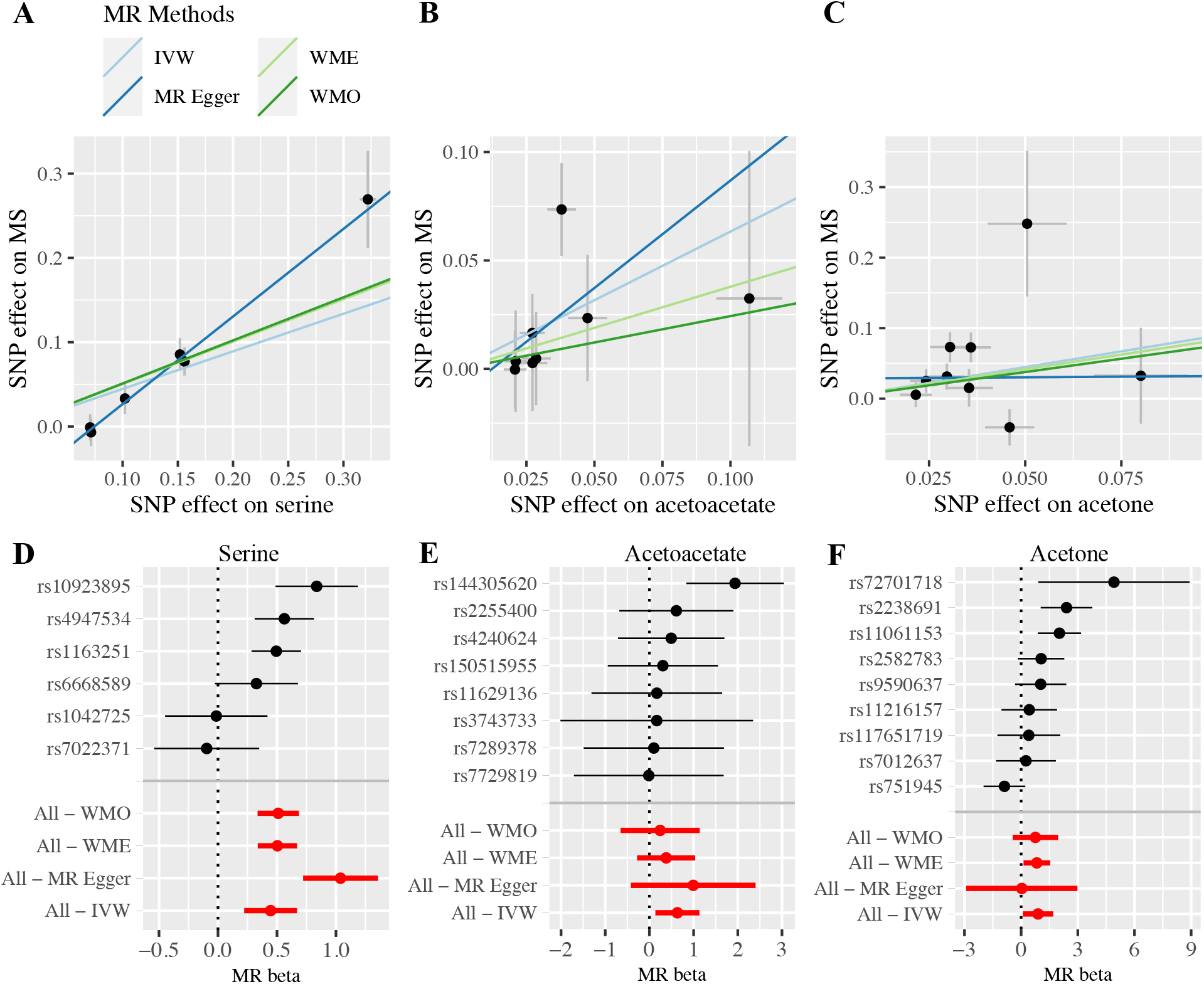
MR estimated effects of three metabolites on MS. Scatter plots for serine (A), acetoacetate (B), and acetone (C) illustrate the individual SNP effects on the metabolite and MS and the estimated linear causal relationship between the metabolite and MS by applying four MR methods. Forest plots for serine (D), acetoacetate (E), and acetone (F) show the causal effect estimates based on individual SNPs and based on all SNPs using four MR methods. MR: Mendelian randomization; MS: multiple sclerosis; IVW: inverse variance-weighted; WME: weighted median; WMO: weighted mode; SNP: single nucleotide polymorphism.

Two of the six consistently significant metabolites are acetoacetate (OR = 2.47, 95% CI = 1.14 – 5.34, *P* = 0.021) and acetone (OR = 2.45, 95% CI = 1.02 – 5.90, *P* = 0.046), both of which are associated with a higher MS risk (Figure 3). Another notable metabolite is uridine, which is positively associated with the MS risk (OR = 1.45, 95% CI = 1.10 – 1.91, *P* = 0.008).

## Discussion

Our metabolome-wide MR study, the first of its kind for MS, prioritized a list of 29 circulating metabolites that likely have causal associations with the risk of MS. Our results highlighted metabolites in lipid metabolism (e.g., cholesterol and phospholipids in large VLDL and very large HDL), amino acid metabolism (e.g., serine and lysine), and energy metabolism (e.g., acetoacetate and acetone).

Altered lipid metabolism is well-known in MS patients^33^. A metabolomics study, comparing relapsing-remitting MS patients (RRMS) to age- and sex-matched healthy volunteers, found that cholesterol, phospholipids, and triglycerides are elevated in the larger subclasses of VLDL and HDL^8^. Two previous MR studies examined the causal roles of HDL-C, low-density lipoprotein cholesterol (LDL-C), and triglycerides in MS. Using GWAS of blood lipids that are independent of our GWAS of metabolomics, they found that HDL-C is positively associated with the MS risk, while no significant effects were found for LDL-C and triglycerides^17, 34^. Our results consistently revealed that total cholesterol, cholesterol esters, and phospholipids in very large HDL are associated with a higher MS risk. Also, we did not find significant effects of lipids in LDL. Our study showed that lipids in large VLDL are associated with a lower MS risk. It is important to note that the previously observed elevated levels of lipids in the larger subclasses of VLDL in RRMS patients may be confounded by reserve causation, as lipid levels may respond to the progression of MS. Our study highlighted the importance of examining the role of lipoprotein subclasses in MS.

Altered circulating levels of amino acids and gamma-glutamyl amino acids have been observed in MS patients^2, 3^. A higher serum level of lysine was observed in MS patients when compared to healthy controls^6^, and also in MS patients who are in relapse in comparison to those that are a few months after the last relapse^35^. The pattern for serine is more complex. A lower plasma concentration of serine was observed in RRMS patients^36^, but a higher serum serine level was found in secondary progressive MS patients^37^. On the other hand, in the experimental allergic encephalomyelitis (EAE) rat model of MS, elevated levels of both lysine and serine were observed in the spinal cord and the brain^38^. Our MR analysis indicates that individuals with genetic capacities for higher lysine and serine are more likely to develop MS. Interestingly, it has been shown that EBV infection, a known risk factor for MS^1^, upregulates the import and biosynthesis of serine in B cells^39^. Moreover, serine is a precursor for phosphatidylserine and sphingomyelin, both of which are key lipids in myelin and implicated in the demyelination process of MS^40, 41^. Therefore, our observations of serine and lipids may be related. As for the two gamma-glutamyl amino acids, both gamma-glutamylleucine and gamma-glutamylphenylalanine were observed to be higher in MS patients than in healthy controls, although only gamma-glutamylleucine reached statistical significance. But both of them were significantly elevated in MS patients receiving vitamin D supplementation^5^. Our MR analysis suggests that gamma-glutamylleucine increases, while gamma-glutamylphenylalanine decreases, the risk of MS. Our results call for future studies into the effects of these amino acids before the onset of MS.

Disrupted nucleotide metabolism and energy metabolism are commonly observed in MS^2, 3^. Consistent with our MR result that individuals with a higher genetic capacity for uridine have a higher MS risk, it has been observed that the serum uridine level is higher in MS patients^7^. Similarly, previous case-control studies observed that MS patients have elevated levels of acetoacetate and acetone in the plasma and the cerebrospinal fluid^4, 42^. Our MR analysis supports the causal roles of these metabolites in the development of MS. Notably, the higher circulating levels of ketone bodies (i.e., acetoacetate and acetone) may reflect a protective shift in energy metabolism in MS patients, and ketogenic diets have shown suggestive benefits for MS patients^43^. It is of great interest to investigate the roles of ketone bodies in the development of MS, in addition to its treatment.

The present study has multiple strengths. First, the two-sample MR study design mitigates biases from residual confounding and reverse causation in observational association studies. Second, we examined an extensive list of metabolites to systematically investigate their causal roles in MR risk. Third, two thresholds (*P* < 5 × 10^−8^ and *P* < 1 × 10^−6^) were applied to select genetic instruments, and results are consistent across the two analyses. All metabolites examined have strong genetic instruments (all F-statistics >10), mitigating possible biases from weak instruments. Fourth, we applied six MR methods to assess the robustness of causal associations and effect directions, including IVW with a multiplicative random-effects model, MR-Egger, WME, WMO, MR-PRESSO, and MR Steiger. Fifth, most of our identified metabolites have been previously associated with MS status or severity in traditional epidemiological studies. One (i.e., HDL-C) has been found in previous MR studies, while the other metabolites are novel findings from our study.

Nonetheless, several limitations should be considered when interpreting our results. First, our study could not completely rule out the possible presence of horizontal pleiotropy, although we performed comprehensive MR analyses to confirm consistent causal estimations. Second, some metabolites in the original metabolomics data were excluded from our analysis due to their lack of three or more genetic instruments. Third, some metabolites were present in two metabolomics GWAS, mainly met-c and met-d, but they were only significant in one MR analysis. The different cohort characteristics, study designs, and sample sizes may be underlying these differences. However, we did find that the MR estimates between the two metabolomics GWAS are highly correlated (Supplementary Figure 1). Fourth, our study could not differentiate between MS subtypes. It is of great interest to perform a similar analysis for MS subtypes in the future when large GWAS of these subtypes become available. Fifth, the current MR methods assume a linear relationship between the exposure and the outcome, which may not be the case for some metabolite and MS. Sixth, the MR estimates reflect the lifelong effects of an exposure and provide no information about the critical window of the exposure action. Last, our study was restricted to individuals of European descent to reduce possible bias from population stratification, but it limits the generalizability of our results to other populations.

Our metabolome-wide MR study prioritized metabolites, such as lysine, serine, acetone, acetoacetate, and various lipids, in the lipid, amino acids and energy metabolism that likely play causal roles in the development of MS. They may serve as diagnostic biomarkers to identify individuals at high risk for early prevention. Future studies on these metabolites will further our understanding of the MS pathogenesis and evaluate the efficacy of these metabolites as therapeutic targets.

## Data Availability

All GWAS summary statistics could be accessed through the OpenGWAS project21, 22. The MR analysis scripts can be found at https://github.com/yitangsun/metabolome-wide-MR-for-MS.

## Authors Contribution

KY perceived and designed the study. AG, YS, and TK performed data analysis and prepared visualizations. YZ and KY interpreted the results. AG, YS, and KY wrote the paper. YZ critically revised the paper. All authors read and approved the final manuscript and took responsibility for the integrity of the work as a whole.

## Supplementary Figures

**Supplementary Figure 1.**
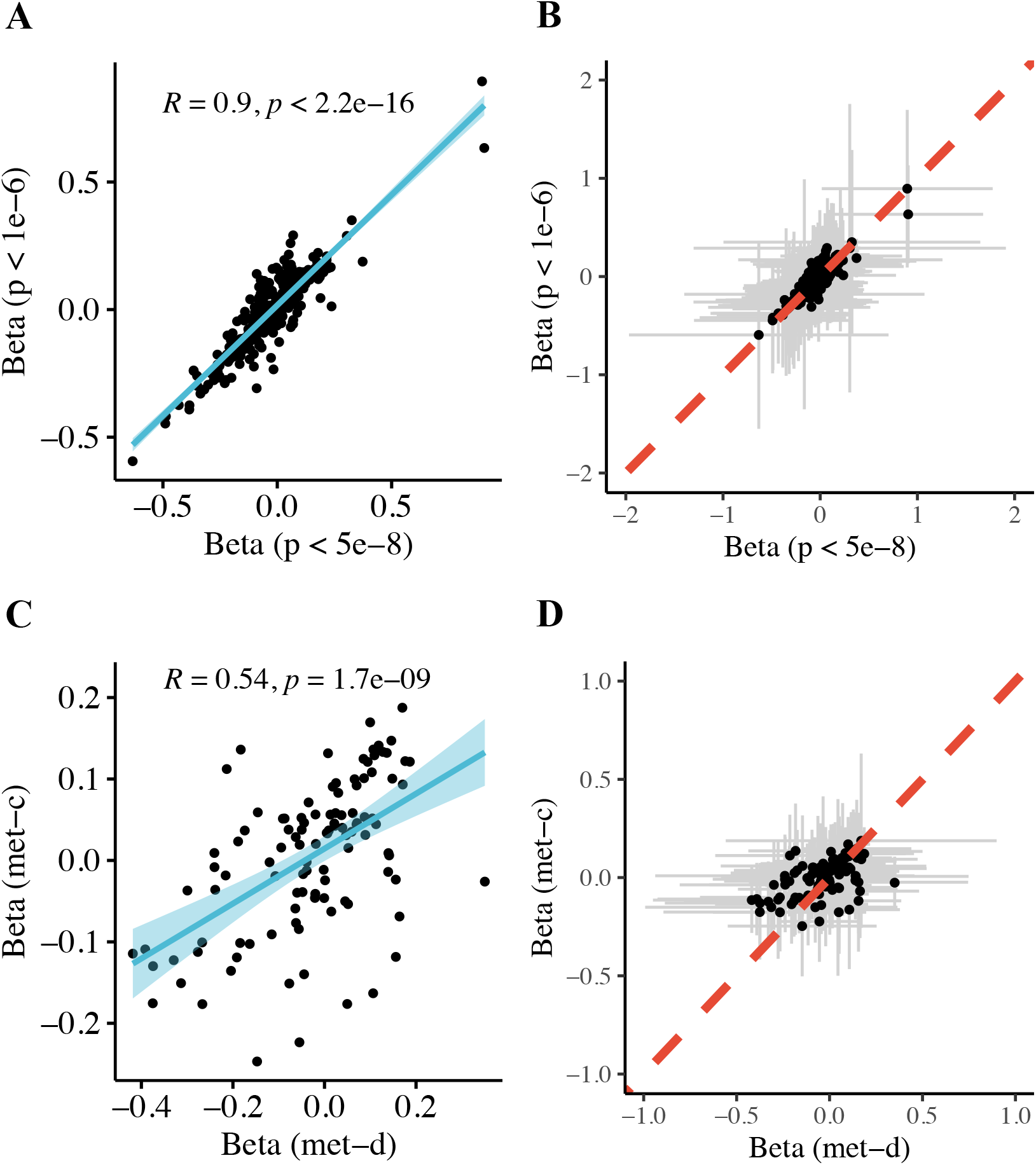
Consistency of MR estimates for the same metabolites between two significant cutoffs, *P* < 5 × 10^−8^ and *P* < 1 × 10^−6^ (A, B), and between two metabolomics GWAS, met-c and met-d (C, D). Only point estimates from the multiplicative random effect IVW method were shown in A and C. The blue line is the linear regression line. The corresponding 95% confidence intervals were shown in B and D. The red dashed line indicates y = x.

## Supplementary Tables

**Supplementary Table 1**. All Mendelian randomization results using the instrumental variables selection threshold (P < 5 × 10^−8^). b: causal effect size; se: standard error; pval: p-value; IVW_MRE: inverse-variance weighted random-effects model; IVW_FE: inverse-variance weighted fixed-effects model; Egger: MR-Egger; Het: heterogeneity; W_Med: weighted median; W_Mod: weighted mode; nsnps: number of SNPs retained for this analysis; SD: standard deviation of metabolites.

**Supplementary Table 2**. All Mendelian randomization results using the instrumental variables selection threshold (P < 1 × 10^−6^). b: causal effect size; se: standard error; pval: p-value; IVW_MRE: inverse-variance weighted random-effects model; IVW_FE: inverse-variance weighted fixed-effects model; Egger: MR-Egger; Het: heterogeneity; W_Med: weighted median; W_Mod: weighted mode; nsnps: number of SNPs retained for this analysis; SD: standard deviation of metabolites.

